# Childhood Behavioral and Mental Health Problems in Jordan

**DOI:** 10.1101/2020.05.26.20113910

**Authors:** Arwa Nasir, Laeth Nasir, Amira Masri

**Author notes:** Corresponding author: Arwa Nasir MBBS, MSc. MPH, Visiting Fulbright Scholar, University of Jordan 2019/2020, Associate Professor, Department of Pediatrics, University of Nebraska Medical Center, 982167 Nebraska Medical Center, Omaha NE 68198-2167, Office.

## Abstract

**Introduction:** Pediatric behavioral and mental health disorders are a leading cause of childhood morbidity worldwide. International and regional data from Arab countries confirm the high prevalence and societal cost of these problems. No data exist that describe the range of childhood behavioral problems encountered by pediatricians in Arab countries.

**Methods:** Qualitative phenomenological methodology was used to collect information. We conducted 8 focus group interviews with physicians in different practice settings across the capital of Jordan. A total of 36 physicians participated in the interviews.

**Results:** Themes that emerged were organized in 4 categories:

1. Specific behavioral disorders: A number of behavioral and mental health disorders were commonly encountered. Autism was reported to be the most commonly encountered severe neurodevelopmental disorder. The scarcity of effective and affordable management and referral options compounded the impact on families and physicians.
2. System related themes: Physicians perceived deficiencies in training, detection and management of behavioral disorders, a lack of mental health services and quality referral options.
3. Family related themes: Several factors were perceived to contribute to behavioral morbidity such as family structure and parenting practices.
4. Sociocultural themes: Societal perceptions that childhood behavioral problems were a family responsibility resulting in reluctance of medicalization or discussion with physicians. Cultural differences in the interpretation and tolerance of certain childhood behaviors rendered ‘standard’ diagnostic criteria problematic.

**Discussion and conclusions:** Pediatricians frequently encounter childhood behavioral problems in clinical practice. Barriers to management of these issues include inadequate training and unique sociocultural, family and system factors. Tailored strategies will need to be developed to improve the detection and management of childhood mental health and behavioral problems in Jordan. Further research to explore, develop and test culturally grounded strategies to address these disorders in Arab countries is needed.

**What is already known?:** Pediatric behavioral problems are antecedents of adult mental health problems. Little is known about pediatric behavioral and mental health problems encountered by pediatricians in Arab countries or the specific barriers to addressing these problems from the perspective of practicing clinicians.

**What are the new findings?:** Jordanian pediatricians encountered a range of pediatric behavioral problems in their clinical practices. The specific factors that contributed to the prevalence of and perceived difficulty in the diagnosis and management of pediatric behavioral and mental health problems were identified. These related to medical systems, family factors and psychosocial factors.

**What do the new findings imply?:** A contextual and holistic understanding of specific problems encountered by pediatricians in practice as well as the barriers to addressing them is critical to the development of culturally informed interventions to detect and effectively manage pediatric behavioral and mental health problems in Arab countries.

## Introduction

The global burden and impact of mental health problems is high and rising. [1] In particular, childhood behavioral disorders are highly prevalent. The WHO estimates that 10-20% of children and adolescents worldwide experience significant mental disorders. [2] These disorders are the leading cause of disability among young people, and are particularly impactful to society because of their negative effects on the developmental trajectory of children.[3] This can negatively affect their future educational attainment, health and productivity.

Additionally, childhood behavioral disorders are often the precursors of adult mental health disorders. [4-8] It is estimated that more than 70% of adult mental health problems begin in childhood or adolescence. [9]

Even in countries in which there are few barriers to mental health diagnoses and treatment, mental health and behavioral disorders that reach the threshold of diagnosis represent the tip of the iceberg.[23] Children with milder forms of behavioral dysfunction often suffer significant morbidity without ever getting the help they need. Untreated, these problems often persist and worsen and present later as harder to treat adult mental health disorders. [9]

In the United States (US) and many other western countries, most mental health and behavioral problems are identified and managed within the health care system. Nearly all children with behavioral or mental health problems present initially to pediatricians or other primary care providers. [24-29] Most of the assessment and management of these problems, including pharmacological treatment, also takes place in the primary care office. Only a minority of children receive specialized psychiatric and mental health services, at least in part because of lack of referral resources. [30,31] A report by the WHO indicates that although pediatricians are recognized as the main mental health providers for children, fewer than 1 in 4 pediatricians receive any mental health training. [32] Studies from the US indicate that behavioral problems represent a significant clinical burden in primary care pediatric practice. Among the reported barriers to effective recognition and management of behavioral and mental health problems are insufficient training, short visit times and low reimbursement. [28,30,33,34]

A range of evidence-based strategies have been shown to be effective in the prevention and treatment of many childhood behavioral and mental health disorders.[10-14] Effective mitigation and treatment of these disorders in childhood limit negative outcomes and abort the progression of these disorders to much more difficult to treat mental health disorders in adults[15] However, it has been noted that the paucity of locally applicable data regarding childhood behavioral and mental health problems from Low and Middle Income Countries, (LMIC) is a major barrier to addressing these disorders. [16]

Culture and society strongly influence the ways in which behavioral disorders are conceptualized, classified and approached.[17,18] Stigma around mental health, community attitudes and support as well as medical system characteristics also contribute to whether and how these conditions are addressed and treated. [19-22]

It has been estimated that 100-140 million people living in Arab countries suffer from one or more mental health disorders.[35] Stigma, lack of services and cultural factors related to disclosure have been identified as major barriers that limit help seeking behavior resulting in under treatment of patients with mental health disorders.[17,22,36-38] In addition, it has been proposed that widespread system barriers exist. These include low prioritization of mental health, a lack of resources and poor organization of mental health services. [39]

Comprehensive epidemiological data of pediatric behavioral problems in Arab countries are lacking. However, the few studies available suggest high rates of specific behavioral disorders. A meta-analysis of studies on Attention Deficit/Hyperactivity Disorder (ADHD) in Arab countries suggested that the prevalence of this condition ranged between 1.6 and 16%.[40] A 2018 paper that studied a sample of adolescent Jordanian students found that 11.7% of these students had psychological and behavioral problems. Most common were anxiety and depression (14%), followed by conduct problems (12.5%) and hyperactivity (7%). The study also found significant correlations between behavioral scores and scores of academic achievement. Another study from Egypt found a high prevalence (34.7%) of children had abnormal scores on a measure of observed problematic behaviors, the Strengths and Difficulties Questionnaire.[41]

A large epidemiological study in the Middle East and North Africa (MENA) region found that families faced with mental health or behavioral issues preferentially turned to other family members, friends and other community resources for advice. Frequently interventions by traditional and/or religious healers were sought for these problems before a small minority turned to the formal medical system. (Usually the primary care physician). [42] Other studies indicate a high burden of mental health problems among children and families seeking care at hospitals and other health care facilities. [43,44] Little is known about the ways in which these patients present to pediatricians in clinical practice, what conditions are most commonly seen, or in what contexts.

This study aimed to explore the range of childhood behavioral health problems in the community as well as the subjective burden that these conditions have on families and pediatricians by examining the experience of pediatricians who serve children and families in the community. Using a qualitative methodology allowed us to explore the observed range of behavioral morbidity, and the ways in which these conditions are conceptualized and approached from the perspective of pediatricians practicing in Jordan. Additionally, distinct psychosocial and cultural formulations and practices could be detected, as well as providing naturalistic observations of the presentations, barriers and treatments encountered in clinical practice.

## Methods

Study design: We used a qualitative phenomenological research methodology to explore the experiences and attitudes of physicians towards behavioral problems and disorders in children.

Study population: Eight focus group interviews were conducted from October 10 until December 30^th^ 2019 with groups of pediatricians in Amman, the capital of Jordan.

Jordan is a Lower Middle-Income Country (LMIC) with a population of 9 million people. About 40% of the population is under 15 years of age.

The pediatrician groups were chosen to represent a variety of practice settings. The focus groups ranged from 1-10 individuals and included 3 groups of private practice community pediatricians, 2 groups of government (Ministry of Health) pediatricians, one group of University-based academic pediatricians, and 2 groups of pediatric house officers in training. The total number of interviewees was 36.

Research procedures: The research protocol was approved by the ethics boards of the University of Nebraska and the University of Jordan. Informed consent was obtained verbally from the interviewees, and they were provided with an information sheet. The interviews lasted between 51 and 75 minutes. The interviews were conducted in the physicians’ workplaces. They were audio recorded, transcribed and translated verbatim by the primary investigator. The primary interview question was: What types of pediatric emotional/behavioral or family and psychosocial problems do you encounter in your practice? Follow up and probe questions were asked as needed. Interviews of groups continued until inductive thematic saturation was achieved.

Data analysis: NVivo 12 software was used to assist with data management and coding. Data were analyzed by the investigators who independently read the transcripts, then met to review each transcript. During these meetings the transcripts were read and coded, using an inductive approach to the development of the themes, and consensus between the three coauthors was achieved. In a few areas of the transcripts there were Arabic words or phrases that could not be directly translated to English. All three coauthors (who are bilingual and culturally experienced) reached consensus on the meaning of these words or phrases.

Patient and Public Involvement: There were no public participation in the design or implementation of this study.

## Results

Several distinct themes around behavioral and mental health disorders were identified by the authors during the analysis. These themes fell into 4 general categories. They were: descriptions of specific behavioral disorders encountered clinically, themes related to the family, themes related to the medical system and sociocultural themes. There were some differences in thematic emphasis in different groups, but there was high level of consistency among all the groups on each of the themes.

### Behavioral and psychosocial conditions encountered by physicians

Table 1 lists all of the specific behavioral and mental health conditions that physicians reported that they had encountered in the clinical setting.

**Table 1:** List of childhood behavioral problems encountered by participants.

| **Condition** |
| --- |
| **Autism** |
| **Screen overuse** |
| **ADHD** |
| **Gender dysphoria** |
| **Substance use/ smoking** |
| **Bullying** |
| **Anger** |
| **Enuresis/Encopresis** |
| **Sleep problems** |
| **Suicide** |
| **Speech delay** |
| **Tics** |
| **Domestic violence** |
| **Child abuse/neglect** |
| **Depression/Anxiety** |
| **Eating disorders** |
| **Medical non-compliance** |
| **Masturbation** |

#### Autism

There was general agreement in all the groups that autism was the most prevalent serious neurobehavioral condition encountered by pediatricians. *I wonder, with as many young children as we see, what happens to them as they get older”?* Physicians in all of the focus groups, especially in the outpatient setting, perceived autism to be common and extremely problematic to manage. It was also perceived to be more common than ADHD. Physicians also reported their perception that the prevalence of autism has increased significantly in recent years. The physicians reported specific difficulties in making the diagnosis and counseling the family. *“I refer all these kids, they need something that I cannot provide for them”*. Physicians had low confidence in their ability to make a diagnosis of autism, which they related to having received little education or training about the condition. They also reported receiving little ongoing education about autism: “*Honestly, it is a very difficult subject, there is Autism, but also there is Autism spectrum! It is confusing”*. Pediatricians reported they referred children to neurologists to establish the diagnosis and rule out other conditions. Neurologists could also prescribe anticonvulsant medications or “psychoactive” medications when needed, which requires a special license not available to generalists: *“the neurologists are carrying the burden of these cases. “*

Lack of referral options for the management of the condition was also problematic. Several physicians stated that they felt that the few available diagnostic and treatment centers were inadequate to meet the needs of patients and were generally unaffordable for average Jordanians.

The combination of large numbers of patients with autism or suspected autism, the lack of available diagnostic and treatment services and the disruptive nature of the disorder on the child and family, and the limited government resources available for this condition resulted in high levels of frustration among physicians and parents around this diagnosis. *“If I had the authority to change things in this country, the first thing I would do would be to try to help these families, they are suffering greatly”*.

#### Screen time overuse

Physicians were unanimous in their perception that the excessive use of screens in young children, particularly mobile phones and tablets was resulting in high rates of speech delay and impairments in social and emotional skills. The pediatricians in our sample considered the condition to be very common at all socioeconomic levels. Many physicians thought that parents, especially mothers, used screen media to manage their children’s behavior while they attended to other tasks. *“I call it ‘mobile autism ‘. You feel the child does not have autism but acts like he does… I think it affects social interaction. The child loses their ability to interact with other people. They lose interest in their surroundings”*.

#### ADHD

This condition was reported to be more of a problem for physicians working in secondary or tertiary care settings, since suspected cases are immediately referred from primary care to higher levels of care. As in autism, physicians indicated they lacked confidence in making the diagnosis. *“As a pediatrician, I do not know how to make the diagnosis of ADHD, it is a big responsibility”*. They also reported a lack of affordable behavioral services resources for diagnosis and management. *“Sometimes the schools send the kids to psychologists to be evaluated, and that is very expensive and a long tedious process”*.

Medication management for ADHD was not favored by patients, and physicians often reported discomfort in prescribing these medications. *“We don’t like medications either” “prescriptions need a special license, stamp”*. Severe cases were usually referred to a neurologist where the diagnosis was made and medications prescribed. Physicians reported their perception that most children with ADHD end up going untreated. *“They do not get treatment, their education suffers, they quit school”*.

#### Suicide

Reports of attempted or completed suicides, although uncommon, were mostly restricted to physicians who worked in the inpatient setting. Physicians reported that poverty and family psychosocial issues were the main causes of this condition. Also authoritarian parenting practices were thought to contribute to suicidal behavior, particularly in female adolescents.

#### Child abuse

Medical neglect was most commonly cited in this category, and was reported to be particularly common in children with special health care needs. Again, it was felt that poverty and socioeconomic factors were primary drivers of this problem.

#### Masturbation

This was mentioned as a problem in two of the focus groups. Although the physicians acknowledged that masturbation is a developmental stage variant, they had difficulty with the management of this condition due to the societal intolerance and sensitivity of this condition and its sexually charged nature. Physicians reported being uncomfortable counseling parents and schools about this condition and were uncertain about how to best manage it.

#### Gender dysphoria

Among the more experienced physicians, there was general agreement that this condition was being seen more commonly than it had been previously. There was disagreement among physicians as to the cause. Some physicians felt strongly that it was due to ‘confusion’ sowed by parents and others because they did not provide their children with clear gender appropriate guidance, such as dress, behavior. This is exemplified by the traditional practice of dressing young boys as girls until 3 years of age to protect them from the “evil eye”. Some physicians felt that this custom contributes to gender identity confusion and mentioned that they advised parents against it. Other physicians were less certain that the cause was environmental. Overall, the focus group participants did not feel that tolerance of the condition was a viable option.

#### Depression and anxiety

There were few mentions of these conditions in primary care. However, it was acknowledged that this was likely because adolescents usually receive their medical care from internists or family doctors rather than pediatricians. This was perceived to be a problem because adult physicians have less awareness of the emotional needs of young adolescents.

#### Feeding and Eating Disorders

Physicians described a number of commonly encountered problems around eating. These included picky and restricted eating patterns, sometimes attributed to or aggravated by coercive feeding practices by the parents in an effort to ensure that the child is healthy. Other problems mentioned related to overeating and the consumption of junk food. This problem was exacerbated by the ready availability of home delivery of fast food and by lax parental control and supervision. Anorexia nervosa and bulimia were perceived to be rare.

### Themes Related to Mental and Behavioral Health Conditions

#### Family themes

Physicians in all groups commented on deficits in parenting. “We *have a twenty-year gap in parenting”* These included lack of consistency in enforcing rules set for children, and problems with hierarchy and authority within families. These resulted in all sorts of misbehavior, lack of respect for authority as well as eating and sleeping problems. These problems were particularly acute in homes with parental conflict or divorce, which many of the participants reported is increasingly frequent. *“We talked about divorce and separation, I see a lot of that”*.

Mothers exhausted by working outside the home, or caring for elderly or disabled family members tend to leave the child in the care of a mobile device, resulting in young children using these devices for long unsupervised hours.

*“…and the woman working outside the home. She gets out of the house in the morning and returns at 5. That affects the children greatly, she leaves them with the maid and the maid does not care.”*

The role of the extended family as a support system for the children and parents on one hand and as a disruptive influence in the family hierarchy on the other, was mentioned in several of the focus groups. For example, living with or close to extended family could result in the children taking advantage of their indulgence to avoid rules, especially pertaining to dietary restrictions in children with chronic diseases such as diabetes. The prominent role of the grandparents especially the paternal grandmother in making decisions, could result in marginalization and disempowerment of the mother. *“…when grandmothers come to the clinic with families, they usually are dominant and they are the decision makers”*.

In addition, new patterns of family formation, that included increased rates of nuclear family living, meant that children tended to have less supervision than previous generations did. *“…there was a role for the extended family, now that role is not there anymore. Grandmas and grandpas and uncles, it used to be that the family lived together and the mother would say something and the grandmother would say something and the uncle gives advice, now everyone is on their own”*.

Participants also perceived that new norms of social behavior and exposure to social media also place significant stresses on families, particularly mothers who are considered primarily responsible for children’s behavior and upbringing.

#### System Related Themes

A major complaint of physicians in the focus groups was that they lacked the knowledge and training to deal with behavioral and psychosocial issues. They also reported competing clinical priorities which prevented them from dealing with the difficult and time-consuming problem of behavioral diagnoses. Some clinicians reported sometimes suspecting the presence of a behavioral problem, but because they felt unable to deal with it, or anticipated a lack of success, they would not address it. They reported that they did not feel that many of the solutions and formulations presented in their pediatric textbooks or resources allowed them to understand and address these issues, and they reported not having had mentoring in approaching psychosocial issues during their training.

Poor access to services for behavioral or mental health were reported due to high cost, poor quality, long waiting times, or all three. There was a perception that behavioral health in children was not prioritized by the system. *“There is a huge shortage in everything, and behavioral health doesn’t get a turn”*.

#### Sociocultural factors

***“All the society’s problems trickle down to the children!”***

Several subthemes were identified in this category. These included:

### Traditional Expectations of mothers

Focus group participants discussed a number of sociocultural factors that they felt contributed to behavioral and mental health problems among the patients that they encountered. One of the issues felt to be important related to the perception of women’s disadvantaged position in society. Women are traditionally expected to shoulder the primary burden of raising children, housekeeping and caregiving for her husband and any elderly or disabled relatives who may live in the home. *“Many fathers feel that the children are the responsibility of the mother. When the father has this idea that the mother has to do everything, everything goes bad*…*”*

Increasingly women work outside the home, often because of economic imperatives. This often results in exhaustion and inability to devote emotional energy to her children’s problems. *“They work and come home tired. And if they have school age children then the mother will have to help with school work. In addition to all the things she has to do. The mother is crushed”*. Mothers also tend to be blamed for any of the child’s negative health outcomes, whether physical or behavioral. Often if the child is born with anomalies or has a behavioral problem, the mother is blamed and may be in danger of being divorced. “*She is in the weaker position in the relationship. There is not much support for her, so even if the father is in the wrong, all blame will go to the mother, I guess this is how it is in our culture”*.

### Social attitudes towards children’s behavior and behavioral problems

Some behaviors that might be labeled as abnormal or problematic among children in other world regions might be normalized, tolerated or ignored by families and society in Jordan.

*“It is surely a matter of definition,…the definition of what is hyperactivity depends on the environment.”*

*“I think in Jordan we don’t pay attention to the behavioral aspects of children’s health. we just ignore it early on”*.

Other types of behavior may be concealed because of stigma “*families are reluctant to bring behavioral problems to the doctors because of stigma. They try to cover it up”*. The communal nature of society in Jordan means that many individual behaviors, illnesses, and practices are widely known and discussed in the community. Behavioral and mental health issues, in particular are subject to much stigma, and this may reflect poorly on the family.

*“When the child has a behavior problem, this will frankly be a disaster for the family, and the mother will be so embarrassed. The child will scandalize the family and they crack down on him which makes things worse”*.

*“I noticed that families are very resistant to the idea that something is wrong with their child”*.

#### Economic Issues

Another theme identified was related to the economic environment. Difficulty making ends meet places families under stress. This reduces the material and emotional resources parents are able to devote to their children, and exacerbates behavioral problems. *“families are stressed, the whole society is stressed”*

### The Built Environment

Some focus group participants surmised that the environment that many families lived in aggravated behavioral and mental health issues among children. In particular, overcrowding and a lack of venues for family recreation and spaces available for children to play were seen as being important contributors to this problem: *“.they live in small apartments and there are no public parks. There is no place for the children to release their energy. They sit in the small apartment on top of each other, which I think adds to the problem.”*

### School Systems

The physicians stated that in Jordan, students in both public and private schools are assigned excessive amounts of homework to keep up with the curriculum. Since Jordanian society places a very high value on educational achievement, mothers face significant pressure to help their children with homework in the evenings. This ‘study time’ at home is usually charged and tense. The interviewees thought that this negatively impacted the parent-child relationship.

*“the child does not learn the stuff at school and the mother has to teach him…. Instead of the mother having time to go to the gym or have time with her friends, she has to teach the kids. It is a stressful thing for the mother and the kids”*.

### Societal change and globalization

Another important theme that emerged was the ways in which culture change, fueled by globalization was changing the behaviors and expectations of children and adolescents. The availability of information, role models and diversity of lifestyles on the Internet and social media was seen as both an opportunity but also as a danger for children. Families and society often felt at a loss on how to best address or confront these changes. This dynamic led to an increase in intergenerational conflict and alienation which further increased stress and reduced healthy coping strategies among families and children: *“Life is getting more complicated, with the Internet and social media and everything, the society has changed.”*

Some examples of changes in outlook included increases in children’s expectations for consumer goods, their attitudes towards communicating and connecting with their parents, and even changes in the foods they ate. Some of the unhealthy eating behaviors that were encountered by the physicians in clinical practice could be directly traced to influences or ideas that children had observed on the Internet or social media.

### Societal attitude toward physicians and modern medicine

Some of the focus group participants discussed parental and community attitudes toward physicians and modern medicine. They suggested that a combination of factors that included a mismatch in expectations, lack of trust, and strong traditional beliefs all resulted in suboptimal communication and treatment of behavioral and mental health problems among children: *“they trust their neighbors and friends more than they trust the doctor.”*

Pediatricians reported that parents rarely present to the physician with an explicitly behavioral complaint. It is usually brought up indirectly or as a secondary issue or it is detected by the physician. *“Most frequently, the family is there because of another problem”*

Medicalization of behavioral and mental health problems may be strongly resisted: *“most people don’t think we are here for behavioral problems. You are here to give them medications and that’s it. If you think about giving behavioral or social advice, they act like they do not want to hear these things from you, no.”*

Family and community attitudes toward certain treatments, such as medications for ADHD may be stigmatized, and so it may be difficult for the physician to recommend or implement certain treatments. The use of traditional and complementary therapies was also a commonly seen by physicians. *“We have to put the evidence in the context of the culture. We can’t really ignore culture or they will not trust you. And they will go to the doctor who will tell them what they want to hear and whose talk is aligned with their beliefs.”*

## Discussion

Our study is the first of its kind in the region. It was carried out in Amman, the capital city of Jordan, among a purposeful sample of pediatricians who practice there. Therefore, their opinions and experiences may be different from other physicians who work and practice in different settings, regions and countries. This study was designed both to develop an understanding of the childhood behavioral problems encountered by pediatricians practicing in the region, and to generate hypotheses that can be tested in subsequent studies.

Our data suggest that pediatricians in Jordan encounter many patients with behavioral problems. However, limited options for referral or treatment, and a lack of consensus around societal standards of behavior and thresholds for medicalization of behavioral problems all appear to contribute to the gap in services for these patients.

Perceived deficits in physician training in mental and behavioral health was another prominent subtheme in the interviews. While physicians reported feeling well prepared to manage biomedical issues, many articulated difficulty and discomfort in approaching mental health and behavioral disorders.

We also found that the pediatricians in our sample suggested that some of the sociocultural beliefs, traditions and practices of patients and families significantly impacted their approach to diagnosis and treatment of patients. In addition, patient perceptions of the role of the doctor in the evaluation and management of behavioral problems varied. This appeared to be at least partly due to the perception that the sphere of influence of medical practice did not include childhood disordered behaviors. This finding is concordant with previous studies in other countries observing that culturally specific narratives defining the cause of a mental health problem is the key determinant in people’s choice of help-seeking pathway [45]

The pediatricians in this study perceived that diagnostic and treatment services for children with the more severe neurobehavioral disorders such as ADHD and autism was particularly deficient. They also indicated a growing trend-some called it an epidemic-of excessive screen use by young children in the general population that impacted their language, social skills and emotional development. This problem was felt to be exacerbated by poor parenting skills, maladaptive family and social dynamics and a lack of awareness about the harmful effects of excessive screen time on children.

## Conclusion

Understanding the lived experiences, perceptions and practices of physicians who encounter children with behavioral and mental health problems is an important starting point in developing models to serve as the basis of interventions that can be implemented in clinical practice and education in the region. These models will also require culture specific data from families and others about contributing factors and mediators of these problems. Further, improving the capacity of frontline physicians to detect and care for patients with behavioral and mental health problems are likely to require the development of tailored approaches to behavioral complaints. Contextual adaptations might need to be made to western mental health models to ensure their effectiveness in the very different sociocultural environment existing in Jordan.

Successful interventions and improved access to childhood behavioral and mental health services will be critical in reducing mental health morbidity in the Middle East North African (MENA) region. Culturally acceptable and effective approaches, especially those that capitalize on the preexisting strengths of the social and cultural environment of families living in the Middle East would significantly help to reduce childhood morbidity and optimize human potential in the region.

## Data Availability

The data in this research include transcripts of focus group interviews and coded remarks. they are available with the corresponding author. however, due to the nature of the data, complete deidentification of the data is difficult if not impossible and sharing them may violate the confidentiality guidelines.

## References

1 Rehm J, Shield KD. Global Burden of Disease and the Impact of Mental and Addictive Disorders. Curr Psychiatry Rep 2019;21:10. doi:10.1007/s11920-019-0997-0

2 Child and Adolescent Mental Health. https://www.who.int/mental_health/maternal-child/child_adolescent/en/

3 Luyckx K, Tildesley EA, Soenens B, et al. Parenting and trajectories of children’s maladaptive behaviors: a 12-year prospective community study. J Clin Child Adolesc Psychol Off J Soc Clin Child Adolesc Psychol Am Psychol Assoc Div 53 2011;40:468–78. doi:10.1080/15374416.2011.563470

4 Briggs-Gowan MJ, Owens PL, Schwab-Stone ME, et al. Persistence of psychiatric disorders in pediatric settings. J Am Acad Child Adolesc Psychiatry 2003;42:1360–9. doi:10.1097/01.CHI.0000084834.67701.8a

5 Fergusson DM, Boden JM, Horwood LJ. Situational and generalised conduct problems and later life outcomes: evidence from a New Zealand birth cohort. J Child Psychol Psychiatry 2009;50:1084–92. doi:10.1111/j.1469-7610.2009.02070.x

6 Fergusson DM, Horwood LJ, Ridder EM. Show me the child at seven: the consequences of conduct problems in childhood for psychosocial functioning in adulthood. J Child Psychol Psychiatry 2005;46:837–49. doi:10.1111/j.1469-7610.2004.00387.x

7 R. R. Althoff, Verhulst FC, Rettew DC, et al. Adult outcomes of childhood dysregulation: a 14-year follow-up study. J Am Acad Child Adolesc Psychiatry 2010;49:1105–16. doi:10.1016/j.jaac.2010.08.006;

8 Reavis JA, Looman J, Franco KA, et al. Adverse childhood experiences and adult criminality: how long must we live before we possess our own lives? Perm J 2013;17:44–8. doi:10.7812/TPP/12-072;

9 Kessler RC, Amminger GP, Aguilar-Gaxiola S, et al. Age of onset of mental disorders: a review of recent literature: Curr Opin Psychiatry 2007;20:359–64. doi:10.1097/YC0.0b013e32816ebc8c

10 Kato N, Yanagawa T, Fujiwara T, et al. Prevalence of Children’s Mental Health Problems and the Effectiveness of Population-Level Family Interventions. J Epidemiol 2015;25:507–16. doi:10.2188/jea.JE20140198

11 Furlong M, McGilloway S, Bywater T, et al. Behavioural and cognitive-behavioural group-based parenting programmes for early-onset conduct problems in children aged 3 to 12 years. Cochrane Database Syst Rev Published Online First: 15 February 2012. doi:10.1002/14651858.CD008225.pub2

12 Gaete J, Sánchez M, Nejaz L, et al. Mental Health Prevention in Preschool Children: study protocol for a feasibility and acceptability randomised controlled trial of a culturally adapted version of the I Can Problem Solve (ICPS) Programme in Chile. Trials 2019;20:158. doi:10.1186/s13063-019-3245-3

13 Cortese S, Adamo N, Del Giovane C, et al. Comparative efficacy and tolerability of medications for attention-deficit hyperactivity disorder in children, adolescents, and adults: a systematic review and network meta-analysis. Lancet Psychiatry 2018;5:727–38. doi:10.1016/S2215-0366(18)30269-4

14 Klasen H, Crombag A-C. What works where? A systematic review of child and adolescent mental health interventions for low and middle income countries. Soc Psychiatry Psychiatr Epidemiol 2013;48:595–611. doi:10.1007/s00127-012-0566-x

15 Wainberg ML, Scorza P, Shultz JM, et al. Challenges and Opportunities in Global Mental Health: a Research-to-Practice Perspective. Curr Psychiatry Rep 2017;19:28. doi: 10.1007/s11920-017-0780-z

16 Patel V, Kieling C, Maulik PK, et al. Improving access to care for children with mental disorders: a global perspective. Arch Dis Child 2013;98:323–7. doi:10.1136/archdischild-2012-302079

17 Fakhr El-Islam M. Arab Culture and Mental Health Care. Transcult Psychiatry 2008;45:671- 82. doi:10.1177/1363461508100788

18 Gopalkrishnan N, Babacan H. Cultural diversity and mental health. Australas Psychiatry 2015;23:6–8. doi:10.1177/1039856215609769

19 Clement S, Schauman O, Graham T, et al. What is the impact of mental health-related stigma on help-seeking? A systematic review of quantitative and qualitative studies. Psychol Med 2015;45:11–27. doi:10.1017/S0033291714000129

20 Dardas LA, Simmons LA. The stigma of mental illness in Arab families: a concept analysis. J Psychiatr Ment Health Nurs 2015;22:668–79. doi:10.1111/jpm.12237

21 Holder SM, Peterson ER, Stephens R, et al. Stigma in Mental Health at the Macro and Micro Levels: Implications for Mental Health Consumers and Professionals. Community Ment Health J 2019;55:369–74. doi:10.1007/s10597-018-0308-y

22 Zolezzi M, Alamri M, Shaar S, et al. Stigma associated with mental illness and its treatment in the Arab culture: A systematic review. Int J Soc Psychiatry 2018;64:597–609. doi: 10.1177/0020764018789200

23 Angold A, Costello EJ, Farmer EM, et al. Impaired but undiagnosed. J Am Acad Child Adolesc Psychiatry 1999;38:129–37. doi:10.1097/00004583-199902000-00011

24 Briggs RD, Stettler EM, Silver EJ, et al. Social-emotional screening for infants and toddlers in primary care. Pediatrics 2012;129:e377–84. doi:10.1542/peds.2010-2211

25 Briggs-Gowan MJ, Horwitz SM, Schwab-Stone ME, et al. Mental health in pediatric settings: distribution of disorders and factors related to service use. J Am Acad Child Adolesc Psychiatry 2000;39:841–9. doi:10.1097/00004583-200007000-00012

26 Burka SD, Van Cleve SN, Shafer S, et al. Integration of Pediatric Mental Health Care: An Evidence-Based Workshop for Primary Care Providers. J Pediatr Health Care Off Publ Natl Assoc Pediatr Nurse Assoc Pract Published Online First: 2013. doi:10.1016/j.pedhc.2012.10.006;

27 Gardner W, Kelleher KJ, Wasserman R, et al. Primary care treatment of pediatric psychosocial problems: A study from pediatric research in office settings and ambulatory sentinel practice network. Pediatrics 2000;106:E44.

28 Heneghan A, Garner AS, Storfer-Isser A, et al. Pediatricians’ role in providing mental health care for children and adolescents: do pediatricians and child and adolescent psychiatrists agree? J Dev Behav Pediatr JDBP 2008;29:262–9. doi:10.1097/DBP.0b013e31817dbd97

29 Kelleher KJ, Campo JV, Gardner WP. Management of pediatric mental disorders in primary care: where are we now and where are we going? Curr Opin Pediatr 2006;18:649–53. doi:10.1097/MOP.0b013e3280106a76

30 Perou R, Bitsko RH, Blumberg SJ, et al. Mental health surveillance among children - United States, 2005–2011. Morb Mortal Wkly Report Surveillance Summ Wash DC 2002 2013;62 Suppl 2:1-35.

31 Kataoka SH, Zhang L, Wells KB. Unmet need for mental health care among U.S. children: variation by ethnicity and insurance status. Am J Psychiatry 2002;159:1548–55.

32 WHO. Atlas of Child and Adolescent Mental Health Resources. 9241563044_eng.pdf;jsessionid=F02E36AACD5FB708DB02704DE72A802A.pdf

33 Nasir A, Watanabe-Galloway S, DiRenzo-Coffey G. Health Services for Behavioral Problems in Pediatric Primary Care. J Behav Health Serv Res 2016;43:396–401. doi:10.1007/s11414-014-9450-7

34 Meadows T, Valleley R, Haack MK, et al. Physician ‘costs’ in providing behavioral health in primary care. Clin Pediatr (Phila) 2011;50:447–55. doi:10.1177/0009922810390676;

35 Osman OT, Nasir L, Mollica RF, et al. Trauma-Informed Care Survey of Psychiatrists and Primary Care Physicians in the Middle East. Prim Care Companion CNS Disord 2017;19. doi: 10.4088/PCC.17m02157

36 Al-Darmaki FR. ATTITUDES TOWARDS SEEKING PROFESSIONAL PSYCHOLOGICAL HELP:WHAT REALLY COUNTS FOR UNITED ARAB EMIRATES UNIVERSITY STUDENTS? Soc Behav Personal Int J 2003;31:497–508. doi:10.2224/sbp.2003.31.5.497

37 Dempster R, Wildman B, Keating A. The role of stigma in parental help-seeking for child behavior problems. J Clin Child Adolesc Psychol Off J Soc Clin Child Adolesc Psychol Am Psychol Assoc Div 53 2013;42:56–67. doi:10.1080/15374416.2012.700504;

38 Karam EG, Karam GE, Farhat C, et al. Determinants of treatment of mental disorders in Lebanon: barriers to treatment and changing patterns of service use. Epidemiol Psychiatr Sci 2019;28:655–61. doi:10.1017/S2045796018000422

39 Maalouf FT, Alamiri B, Atweh S, et al. Mental health research in the Arab region: challenges and call for action. Lancet Psychiatry 2019;6:961–6. doi:10.1016/S2215-0366(19)30124-5

40 Alhraiwil N, Ali A, Househ M, et al. Systematic review of the epidemiology of attention deficit hyperactivity disorder in Arab countries. Neurosciences 2015;20:137–44. doi:10.17712/nsj.2015.2.20140678

41 Elhamid AA, Howe A, Reading R. Prevalence of emotional and behavioural problems among 6–12 year old children in Egypt. Soc Psychiatry Psychiatr Epidemiol 2009;44:8–14. doi:10.1007/s00127-008-0394-1

42 Karam EG, Mneimneh ZN, Karam AN, et al. Prevalence and treatment of mental disorders in Lebanon: a national epidemiological survey. Lancet Lond Engl 2006;367:1000–6. doi:10.1016/S0140-6736(06)68427-4

43 Al-Sharbati MM, Al-Farsi YM, Al-Sharbati ZM, et al. Profile of Mental and Behavioral Disorders Among Preschoolers in a Tertiary Care Hospital in Oman: A Retrospective Study. Oman Med J 2016;31:357–64. doi:10.5001/omj.2016.71

44 Wang PS, Aguilar-Gaxiola S, Alonso J, et al. Use of mental health services for anxiety, mood, and substance disorders in 17 countries in the WHO world mental health surveys. Lancet Lond Engl 2007;370:841–50. doi:10.1016/S0140-6736(07)61414-7

45 Cauce AM, Domenech-Rodríguez M, Paradise M, et al. Cultural and contextual influences in mental health help seeking: A focus on ethnic minority youth. J Consult Clin Psychol 2002;70:44–55. doi:10.1037/0022-006X.70.1.44

